# Unrelated Stem Cell Donor HLA Match Likelihoods in the US Registry Incorporating HLA-DPB1 Permissive Mismatching

**DOI:** 10.1101/2022.11.14.22282311

**Authors:** Loren Gragert, Stephen Spellman, Bronwen Shaw, Martin Maiers

**Author notes:** **Corresponding Author:** Martin Maiers, 500 N. 5^th^ St., Minneapolis, MN 55401.

## Abstract

Donor-recipient HLA matching at the DPB1 locus improves the outcomes of hematopoietic cell transplantation (HCT). Retrospective outcomes studies found that in transplants matched for all 8 alleles of the A, B, C, and DRB1 loci at high resolution (8-of-8 match), few transplants were also allele-matched at the DPB1 locus. DPB1 allele matching was thought to be logistically impractical, however a DPB1-permissive mismatch model based on T-cell epitope (TCE) reactivity expands the proportion of suitable donors. To understand the likelihood of success for finding a DPB1-permissive donor, we sought to expand population genetic match likelihood models for the US unrelated donor registry, National Marrow Donor Program (NMDP). After extending HLA haplotype frequency estimates to include the DPB1 locus, our models found that the likelihood of having a DPB1-permissive donor was not much lower than the 8-of-8 match likelihoods. A maximum of 5 additional donors would need to be typed to find a more optimal DPB1-permissive donor at least 90% of the time. Linkage disequilibrium patterns between the DPB1 locus and other classical HLA loci varied markedly by haplotype and population, indicating that the known recombination hotspot between DQ and DP gene complexes has not had uniform impact, thus DPB1-permissive donors are easier to identify within minority populations. DPB1 TCE categories were highly predictable from HLA typing at other loci when imputed with extended haplotype frequency data. Our overall results indicate that registry search strategies that seek a more optimally matched HCT donor encompassing HLA-DPB1 permissibility are likely to be highly productive.

**KEY POINTS:**

- HLA-DPB1 permissive donors are available for nearly all patients that have an 8/8 allele match
- DPB1-permissive donors will be easier to identify among 8/8 allele matches for minority patients relative to White patients

## INTRODUCTION

Donor-recipient matching for HLA alleles improves outcomes in hematopoietic cell transplantation (HCT)^1^. Retrospective outcomes studies enabled by observational databases of donor-recipient pairs have over time systematically uncovered the importance of matching at high resolution and for matching at additional HLA loci^2,3^. Outcomes registry studies have found strong evidence for a role of DPB1 matching in transplant outcomes^4,5^.

Identification of HLA-matched donors in large registries is often challenged by incomplete and ambiguous HLA genotyping for clinically relevant loci^6,7^. Over time HLA typing of new recruits for the US National Marrow Donor Program (NMDP) Be The Match Registry has expanded in locus coverage and resolution^8,9^. High resolution DPB1 locus typing has been included at recruitment since 2015, enabling models of HLA match likelihoods to now incorporate the DPB1 locus.

Matching for the majority of the HLA loci has been considered at the high resolution allele level, however driven by the low probability of finding a donor who is HLA-DPB1 allele matched in addition to 8/8 matched at the A, B, C, and DRB1 loci, alternative functional models of DPB1 permissibility were sought that would provide good outcomes without exact DPB1 allele matching.

Two paradigms have emerged for identifying permissive DPB1 mismatches based on predicted immunogenicity, relying on categorizing DPB1 alleles either by protein expression level^10^ or by functional T-cell epitope (TCE) category^11–14^. Regardless of paradigm, non-permissive mismatching for DPB1 both increases the risk of acute graft-versus-host-disease (aGvHD) and reduces the risk of relapse. The TCE paradigm developed by Fleischhauer and colleagues used functional assays to measure DPB1 mismatch immunogenicity, leading to definition of groups of T-cell epitopes on DPB1 molecules based on variants in the coding sequences^14^. In this study we modeled the TCE paradigm, as DPB1 expression levels are currently indeterminate for some DPB1 alleles.

Because of a reported recombination hotspot between the HLA-DQ locus and HLA-DP locus^15^, it has long been assumed that typing at other loci could not practically inform the identity of the HLA-DPB1 alleles. When examining retrospective cohorts that were high resolution matched at A, B, C, and DRB1, most transplant pairs were mismatched at DPB1^1^. However, for most of these cases, a DPB1 match was not sought. There have been some hints in the literature that HLA-DPB1 alleles could be predicted from HLA typing at other loci. A report on HLA disease associations by Okada et al. included visualizations that indicated that linkage disequilibrium values between HLA-A and HLA-DRB1 variants have the same magnitude as those between HLA-DRB1 and HLA-DPB1^16^.

While recruitment HLA typing has become more comprehensive over time, currently only 34% of the donors in the US registry are typed at the DPB1 locus, thus there is a risk that these better matched donors may remain unidentified^7^. A typing study was conducted by NMDP to assess the likelihood of finding a DPB1 match or permissive mismatch^17^, finding an 11% increase in DPB1 permissibility for the best matched donor (69% to 80%) could be achieved with a median of 4 donors typed.

Predictive matching algorithms such as HapLogic have been developed to prioritize confirmatory typing of potentially-matched donors with incomplete or ambiguous HLA typing^6^. HapLogic utilizes US population haplotype frequencies^18^ to make predictions of high resolution allele matching at the A, B, C, DRB1 and DQB1, even when those loci are untyped. However, HapLogic does yet not make DPB1 match predictions. While DPB1 epitope status is provided when donors are high resolution typed for DPB1, there are no probabilistic estimates to guide their search when the typing is missing or ambiguous. Therefore, confirmatory typing at the DPB1 typing locus had not been informed by immunogenetics reference data until 2021 when NMDP released a prototype tool for DPB1 match predictions^19^.

In this study we present HLA match likelihoods for patients searching the US unrelated stem cell donor registry for a more optimal donor with A, B, C, DRB1 high resolution allele matching and TCE permissive mismatching at DPB1. We also sought to analyze linkage disequilibrium patterns between HLA loci to understand why DPB1 TCE categories are often highly predictable when the other HLA loci are typed^19^. We hypothesized that if tools for prioritizing donors for additional typing based on likelihood of match at DPB1 were available, better results could be achieved with reduced time and cost.

## METHODS

### Study Population

HLA genotyping data for the A, B, C, DRB1, DRB3/4/5, DQB1, and DPB1 loci on volunteer registry members was extracted from the National Marrow Donor Program registry as of 2020-06-01. Race/ethnicity categories were self-identified in the NMDP donor recruitment questionnaire. US race/ethnic categories derived from the questionnaire categories used in the analysis are listed in Table 1. All US registry members who were HLA typed for at least the A, B, DRB1 loci using DNA-based methods were included in the genotyping dataset used for estimating HLA frequencies.

**Table 1.** Detailed population categories used operationally for modeling the National Marrow Donor Program (NMDP) / Be The Match registry. The NMDP donor recruitment form captures self-identification of race and ethnicity. Each detailed population category has a corresponding broad population category for summary purposes (AFA – African American, API – Asian or Pacific Islander, CAU – European White, HIS – Hispanic, NAM – Native American).

| **Detailed Population Category** | **Race/Ethnic Description** | **Broad Population Category** |
| --- | --- | --- |
| AAFA | African American | AFA |
| AFB | African | AFA |
| AINDI | South Asian Indian | API |
| AISC | American Indian – South or Central Am. | NAM |
| ALANAM | Alaska Native or Aleut | NAM |
| AMIND | North American Indian | NAM |
| CARB | Caribbean Black | AFA |
| CARHIS | Caribbean Hispanic | HIS |
| CARIBI | Caribbean Indian | NAM |
| EURCAU | European Caucasian | CAU |
| HAWI | Hawaiian or other Pacific Islander | API |
| JAPI | Japanese | API |
| KORI | Korean | API |
| MENAF | Middle Eastern / North African | CAU |
| MSWHIS | Mexican or Chicano | HIS |
| NCHI | Chinese | API |
| SCAHIS | Hispanic – South or Central American | HIS |
| SCAMB | Black – South or Central American | AFA |
| SCSEAI | Southeast Asian | API |
| VIET | Vietnamese | API |

### 7-Locus HLA Haplotype Frequency Estimation

7-locus HLA haplotype frequencies were estimated from HLA genotyping data using an expectation-maximization (EM) algorithm implementation that resolves both allelic and phase ambiguity^20^. Details of the 6-locus version of this EM algorithm (which did not include the DPB1 locus) have been previously described^18^. Briefly, haplotypes were estimated by EM in 2-locus blocks using a partition-ligation approach^21^, with the only key difference in methods was that a HLA Class II haplotype block including DPB1 (DRB3/4/5∼DRB1∼DQB1∼DPB1) was ligated to a HLA Class I block (A∼C∼B) to generate 7-locus haplotype frequencies. To measure similarity in 6-locus frequency distributions with the previous published haplotype frequency dataset, we used a normalized similarity index (I_f_)^22^.

### US Registry Match Likelihood Modeling including DPB1

We extended previous models for 8-of-8 and 7-of-8 allele matching at A, B, C, DRB1 loci (4-locus) to 10-of-10 and 9-of-10 allele matching at A, B, C, DRB1, DQB1 and further extended to include the DPB1 locus at high resolution^23,24^, making the models 12-of-12 and 11-of-12. We implemented the HLA-DP T-cell epitope permissibility paradigm (8-of-8 DP-TCE+ and 7-of-8 DP-TCE+) using methods described in detail in the Supplement. Current US clinical matching guidance does not call for HLA matching at the DQB1 locus^25^, however we also modeled 10-of-10 DP-TCE+ and 9-of-10 DP-TCE+ model for centers that prefer to match at this additional locus based on more recent evidence^26^. The models used NMDP registry sizes for actively listed US adult donors and cord blood units at the end of 2020. The NMDP/Be The Match Registry database includes 8,910,469 US donors along with 14,404,303 non-US donors from organizations that list their donors with NMDP so that they will appear in the upfront search list (Supplemental Table 1). Donor availability for adult donors and cell dose requirements for cord blood transplants that each limit effective registry size were modeled as previously described^24^.

### Modeling Productivity of Ordering Confirmatory DPB1 Typing on Potential Donors

To model donor registry search productivity, we simulated what would be expected if the HapLogic registry matching algorithm^6^ could use linkage disequilibrium information to predict which DPB1 alleles and allele groups would be present in donors that were known be allele-level matched at the A, B, C, DRB1 loci, but untyped at the DPB1 locus. Based on the donor-recipient match probabilities at the DPB1 locus, we estimated the average number of donors needed to type before a DP-TCE permissive donor would be found. Under this scenario, all donors with the same race/ethnicity would have identical DPB1 match probabilities if all other loci were allele-matched, therefore the goal of the analysis was to provide guidance for how many donor DPB1 confirmatory typings should be ordered by the transplant center.

### Measuring Linkage Disequilibrium between HLA Loci and Haplotype Blocks

Based on our modeling results, we hypothesized that differences in linkage disequilibrium values between the DPB1 locus and other loci would explain differences in search productivity among populations. To measure linkage disequilibrium between HLA loci, we used an asymmetric linkage disequilibrium (ALD) metric developed by Thomson and Single^27^. We first calculated pairwise ALD between all individual loci.

Next, we sought to measure the relative impact of the recombination hotspot in HLA Class II between the DQ and DP gene complexes to the level of historical recombination in the rest of the HLA region. Across populations we compared the predictability of the DPB1 locus from the rest of the extended haplotype (A-thru-DQB1) to the predictability of the A locus from the rest of the extended haplotype (C-thru-DPB1) using ALD analysis that treated the extended haplotypes as a locus.

## RESULTS

### High Resolution HLA Haplotype Frequency Distributions Incorporating the DPB1 Locus

Seven-locus high resolution HLA A∼C∼B∼DRB3/4/5∼DRB1∼DQB1∼DPB1 haplotype and allele frequencies for 21 US detailed and five broad populations were computed using an expectation-maximization (EM) algorithm. For each population, the sample sizes and number of individuals with HLA typing at each locus are provided in Table 2. The inclusion criteria is all US donors in the NMDP/Be The Match Registry that were genotyped for HLA-A, -B and -DRB1 (at a minimum) using DNA-based methods. Detailed population categories were used for modeling match likelihoods. Complete haplotype frequency distributions are available as comma-separated value (CSV) files at http://doi.org/10.5281/zenodo.4474442.

**Table 2.**
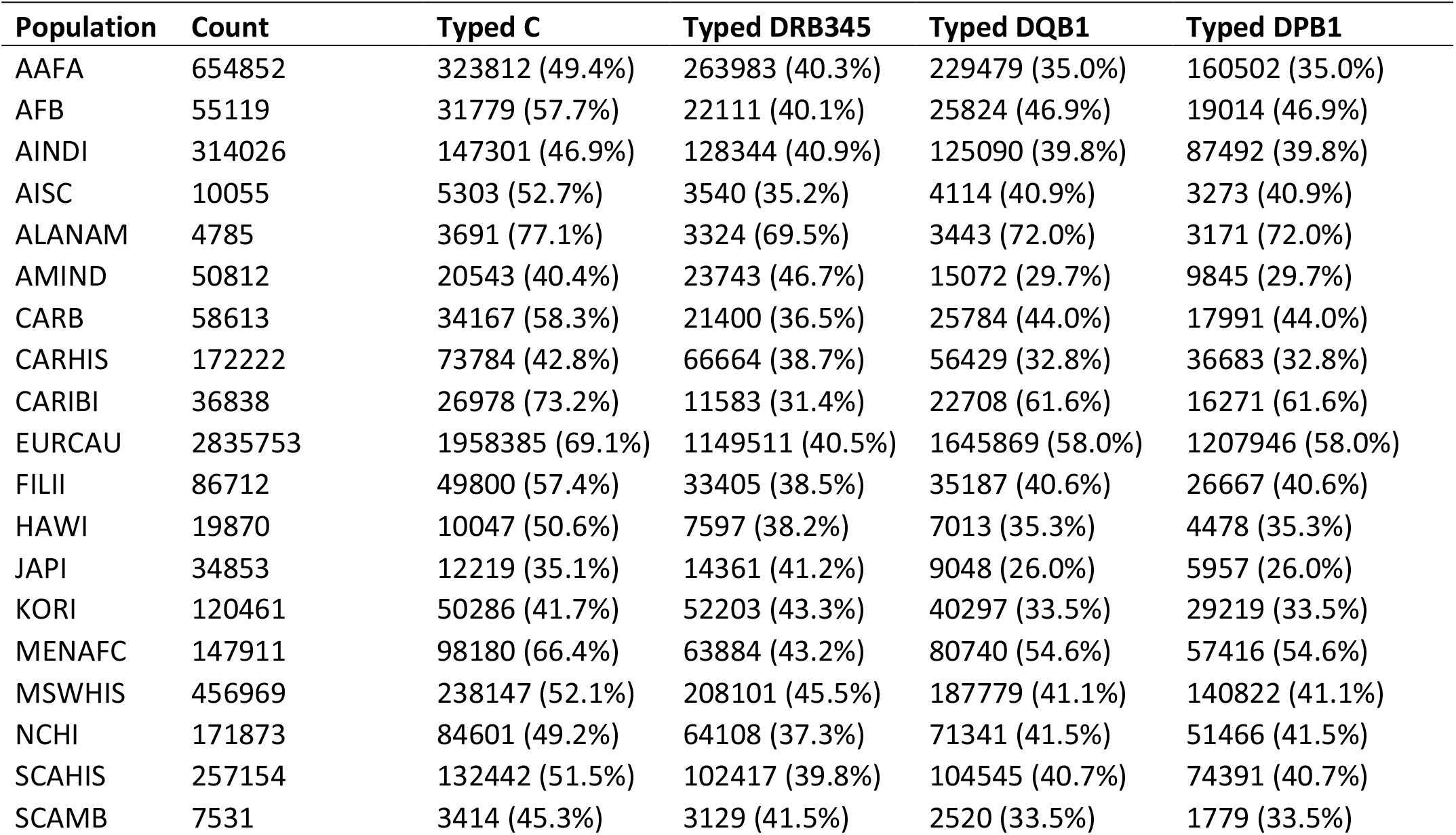
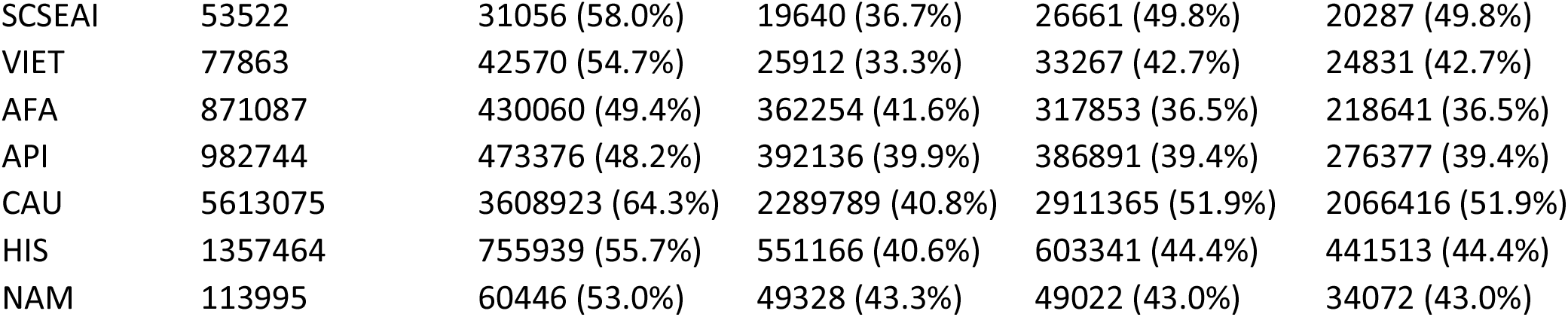
Sample size and composition of donor registry HLA typing data used to estimate 7-locus HLA haplotype frequency distributions, by US population category. The “Count” column indicates the number of donors and cord blood units typed minimally at HLA-A, B, DRB1 loci, and the “Typed C”, “Typed DQB1”; “Typed DRB345”, and “Typed DPB1” columns indicate which subjects are also typed at those loci. The 21 detailed population datasets used for match likelihood modeling are listed first. Data from the 5 broad population categories that follow were used for illustrating differences in allele and haplotype frequencies between populations.

### Current US Registry HLA Match Likelihoods including DPB1 TCE Permissibility

Our models estimate that White Europeans have the best likelihood of having an 8of8 match with DP-TCE permissibility (73.9%), while African Americans have the lowest (24.9%) (Figure 1A).

**Figure 1A.**
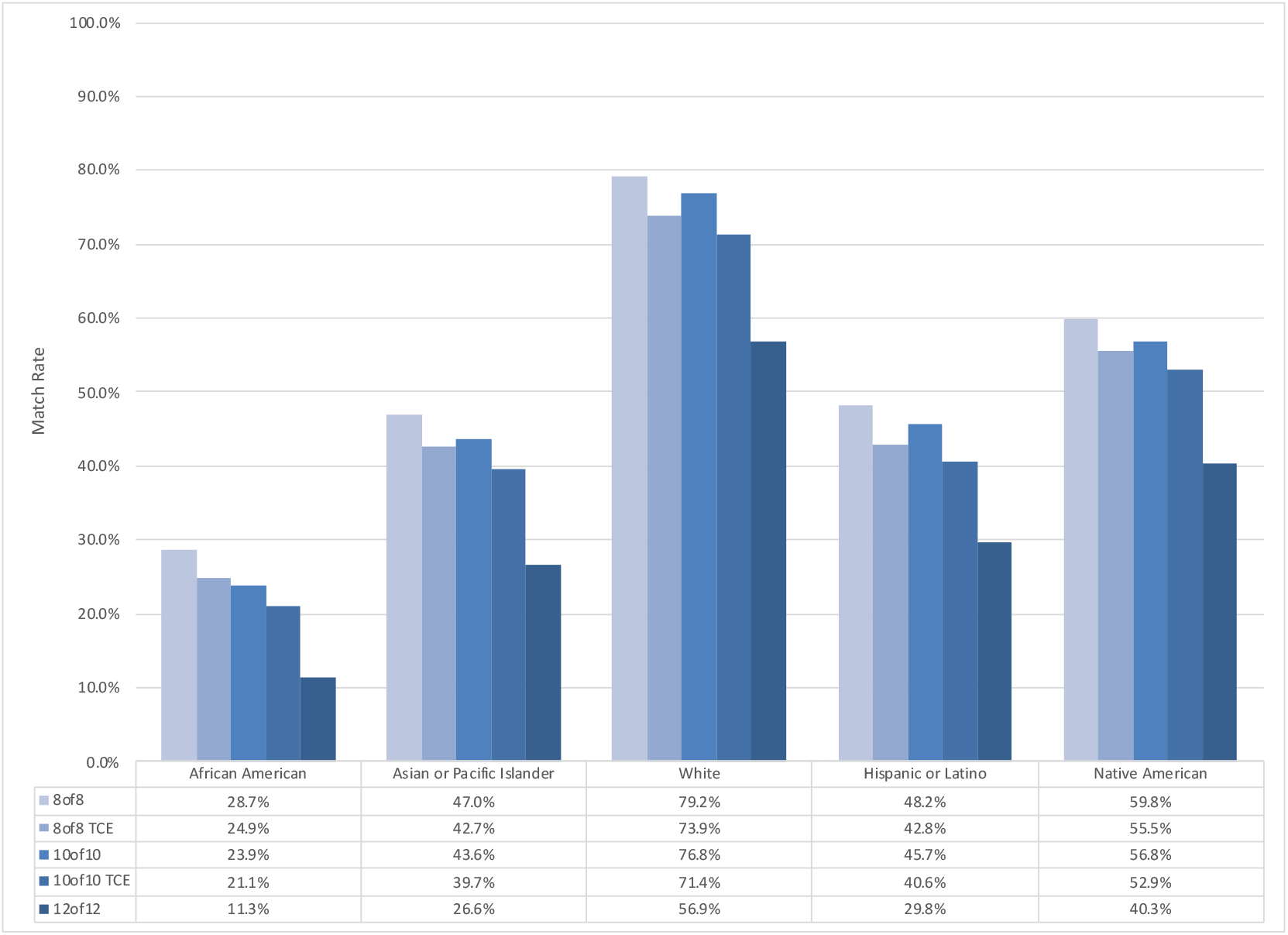
Adult donor HLA match likelihoods for the US registry among 5 broad population categories as of end of 2020. The **“**8of8” match stringency requires high resolution allele matching at 4 of the HLA loci (A, B, C, DRB1). The “8of8 TCE” match stringency requires the donor to be additionally DPB1 T-Cell Epitope (TCE) permissive or DPB1 high resolution allele matched. The “10of10” match stringency requires high resolution allele matching at 5 of the HLA loci (A, B, C, DRB1, DQB1). The “10of10 TCE” match stringency requires the donor to be additionally DPB1 TCE permissive or DPB1 high resolution allele matched. The “12of12” match stringency requires high resolution allele matching at all 6 of the HLA loci (A, B, C, DRB1, DQB1, DPB1).

Conditional on patients already having an 8of8 match, on average, 90.5% will also have a DP-TCE permissive donor. White Europeans patients have the highest conditional likelihood of also having a DP-TCE permissive donor (93.3%), while African American patients are the least likely (86.9%).

Allowing up to one mismatch, the 7of8 DP-TCE permissive match rate ranged from 97% in White European to 77.5% in African Americans. On average, 96.0% of patients who have a 7/8 matched donor will also have an available 7of8 DP-TCE permissively matched donor (Figure 1B).

**Figure 1B.**
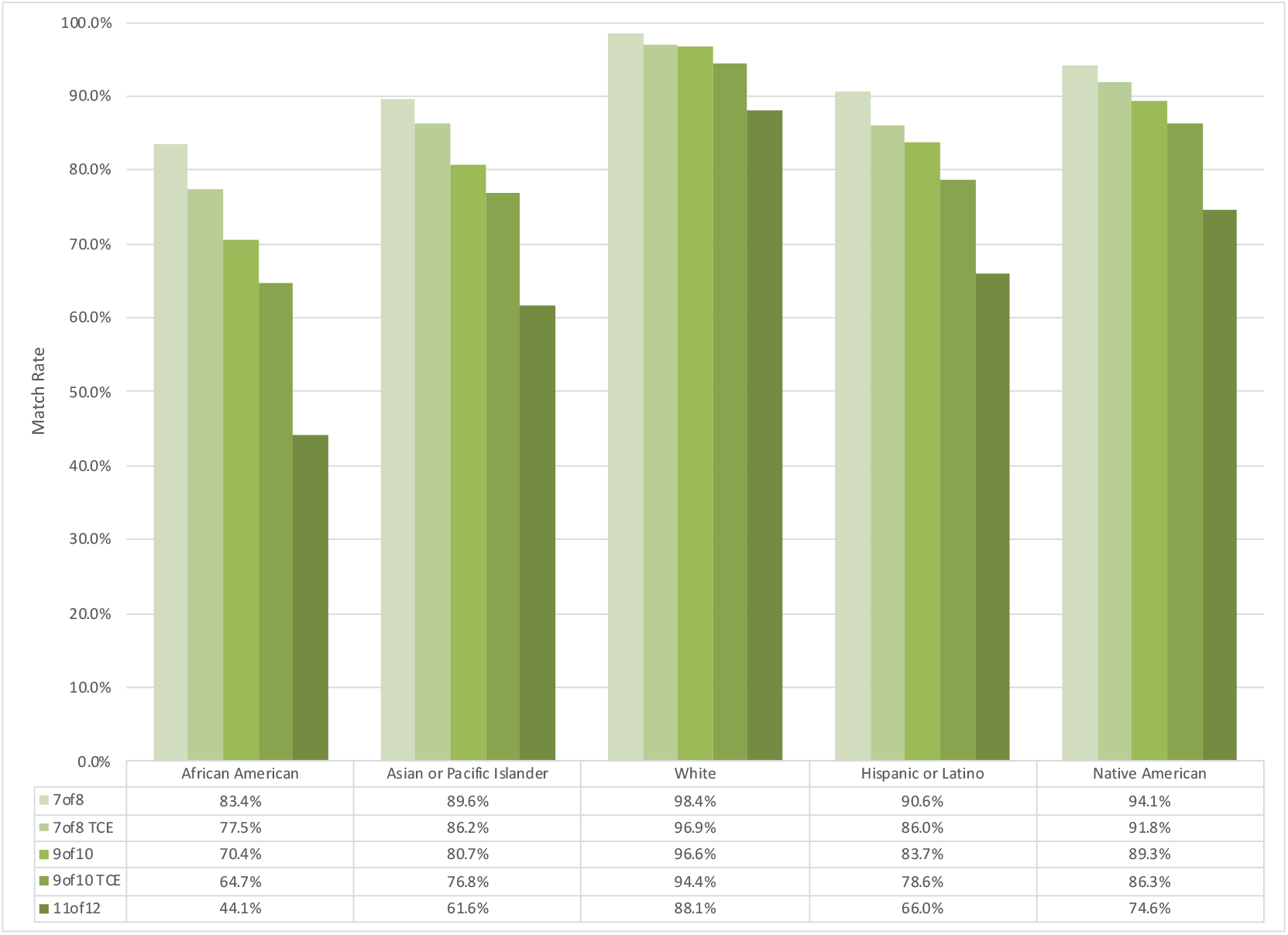
Adult donor HLA match likelihoods for the US registry among 5 broad population categories as of end of 2020, allowing for one mismatch at any locus. The **“**7of8” match stringency allows for up to one high resolution allele mismatch at 4 of the HLA loci (A, B, C, DRB1). The “7of8 TCE” match stringency additionally considers matching at DPB1 T-Cell Epitope (TCE) permissive or DPB1 high resolution allele level, allowing for up to one mismatch. The “9of10” match stringency allows for up to one high resolution allele mismatch at 5 of the HLA loci (A, B, C, DRB1, DQB1). The “9of10 TCE” match stringency additionally considers matching at DPB1 TCE permissive or DPB1 high resolution allele, allowing for up to one mismatch. The “12of12” match stringency allows for up to one high resolution allele mismatch at all 6 the HLA loci (A, B, C, DRB1, DQB1, DPB1).

For centers that consider matching at the DQB1 locus in their protocols, we also provided match likelihoods for the 9of10 and 10of10 stringencies with and without DP TCE. The 10of10 with DP-TCE permissive match rate ranged from 71.4% for White Europeans to 21.1% for African Americans (Figure 1A). The 9of10 with DP-TCE permissive match rate ranged from 94.4% for White Europeans to 64.7% for African Americans (Figure 1B).

Finally, match likelihoods for the 12of12 match stringency show the likelihood of having a high-resolution matched donor across the six loci A, B, C, DRB1, DQB1, DPB1. The 12of12 match rate ranged from 56.9% for White Europeans to 11.3% for African Americans (Figure 1A). The 11of12 match rate ranged from 88.1% for White Europeans to 44.1% for African Americans (Figure 1B).

### Historical US Registry HLA Match Likelihoods including DPB1 TCE Permissibility

To show how donor recruitment and integration of international registries into the NMDP registry has impacted match likelihoods over time, we ran our population genetic models using the registry composition at yearly intervals back to the inception of the US registry. Historical match likelihoods from 1987 to 2020 show match likelihoods for the 7of8, 7of8 TCE, 8/8, 8/8 TCE, and 12/12 stringency. For each match stringency, the match likelihoods for patients showed the same pattern of early rapid improvement through the 1990s, followed by a period of slower but continued improvement until today (Supplementary Figure 1). Diminishing returns on match likelihoods are due to newly recruited donors becoming more and more likely to have the same HLA genotypes represented by previous recruits. Between 1994 and 2000, 2,704,326 donors were added to the registry resulting in a 10% increase 8-of-8 match likelihoods for the White population, however the next 10% increase in match likelihoods between 2000 and 2015 required 9,633,049 additional donors to achieve. The size of the NMDP/Be The Match Registry at year end is provided in Supplemental Table 1.

### DPB1 T-Cell Epitope (TCE) Group Frequencies

Grouping DPB1 alleles by DP-TCE status, TCE Group 3 was by far the most common in every population at over 80% frequency (Figure 3). The TCE group frequencies for the 21 detailed race/ethnic groups are available in Supplementary Methods Table 1. TCE Group 2 was the second most common in all populations except African Americans and Japanese, with the frequency ranging from 19% in Vietnamese to 6% in Filipino.

### Number of Donors Needed to Type at DPB1 to Identify a DPB1 TCE Permissive Match

Patients that had multiple available 8/8-matched donors lacking DPB1 typing had varying likelihoods of finding a DPB1-permissive donor depending on their race/ethnicity and their multi-locus HLA genotype. A randomly selected patient with multiple 8/8 matched donors available will find a DP-TCE permissive match over 70% of the time when one donor is typed, however regardless of population this probability always reached 90% when five donors are typed (Figure 3). Because of population-specific linkage disequilibrium patterns, probabilities varied by population. Strikingly, the White European population had the lowest probability of having a DP-TCE permissive match after typing additional donors. Among detailed population categories, the Filipino population had the highest observed likelihood of finding a DPB1 TCE permissive match after typing one donor (at 91%), while the European White population had the lowest likelihood (at 68%).

### Differences in Linkage Disequilibrium Patterns Between HLA Loci Across Populations Explain Lower Productivity for DPB1 Matching in White Europeans

Linkage disequilibrium patterns differed in magnitude across populations. Figure 4 depicts a heatmap of ALD values between each pair of HLA loci for five broad US population categories. Between White Europeans and African American, the predictability of DPB1 alleles conditional on knowledge of the DRB1 allele was similar (ALD value of 0.271 versus 0.268 respectively), however the predictability of A alleles from DRB1 was much higher in White Europeans than in African Americans (ALD 0.275 versus 0.154).

Modeling linkage disequilibrium in the context of predictive match algorithms that utilize extended haplotype data to predict locus typing, Figure 5 shows that DPB1 alleles are much less predictable in White Europeans when typing at this locus is missing compared with other populations, explaining the lower search productivity found when typing additional donors, as depicted in Figure 2.

**Figure 2.**
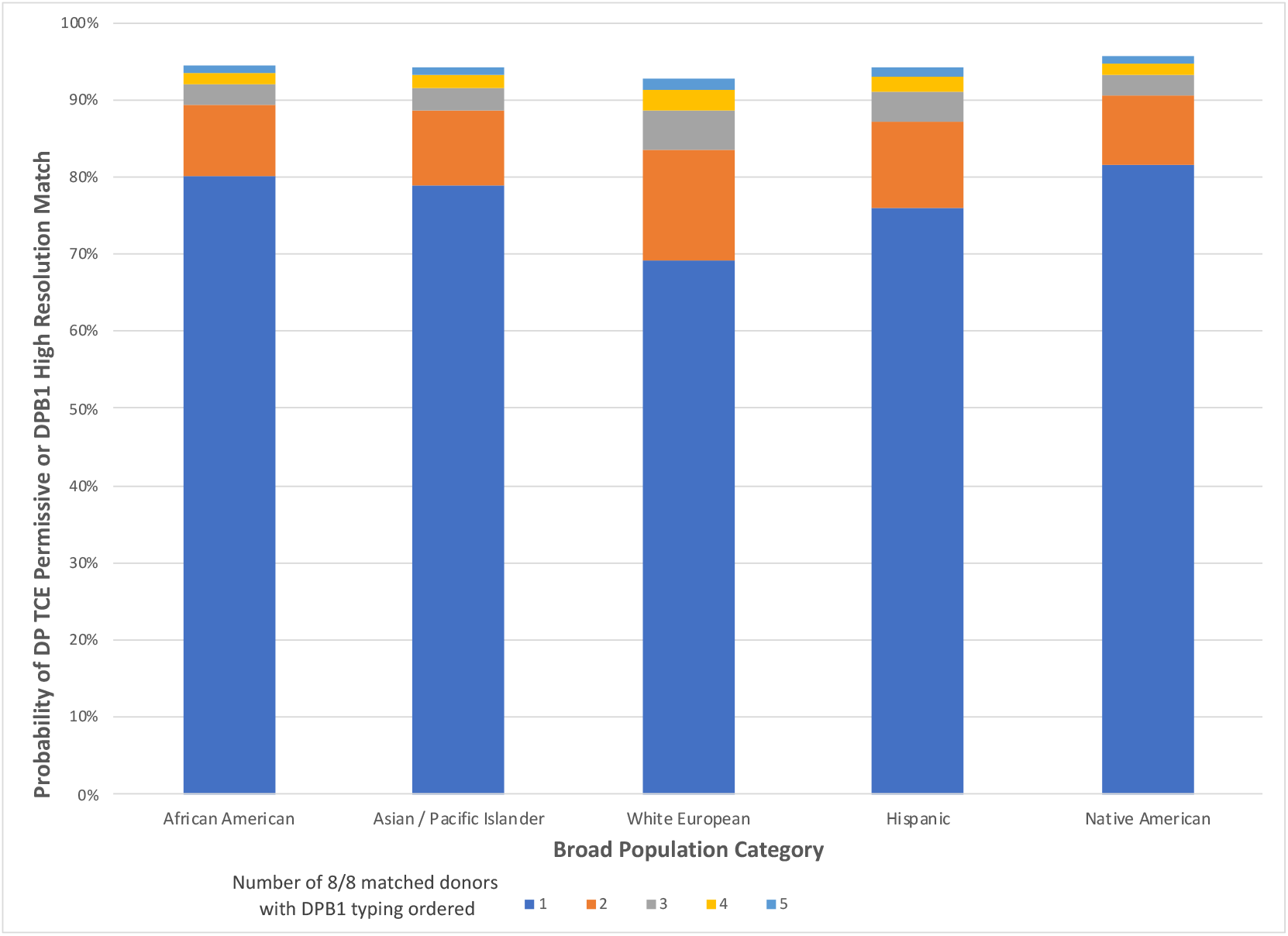
Number of 8/8 Matched Donors to Type at DPB1 Locus to Identify at least one DP TCE Permissive Match, when Donor DPB1 is Untyped. In this scenario, a patient with multiple 8/8 available matched donors with missing DPB1 typing can have up to 5 of the donors typed at the DPB1 locus with the goal of an identifying at least one DPB1 TCE permissive mismatched or high resolution allele matched donor. Results are averages for each population, however search prognosis will differ depending on the patient’s A, B, C, DRB1 genotype.

**Figure 3.**
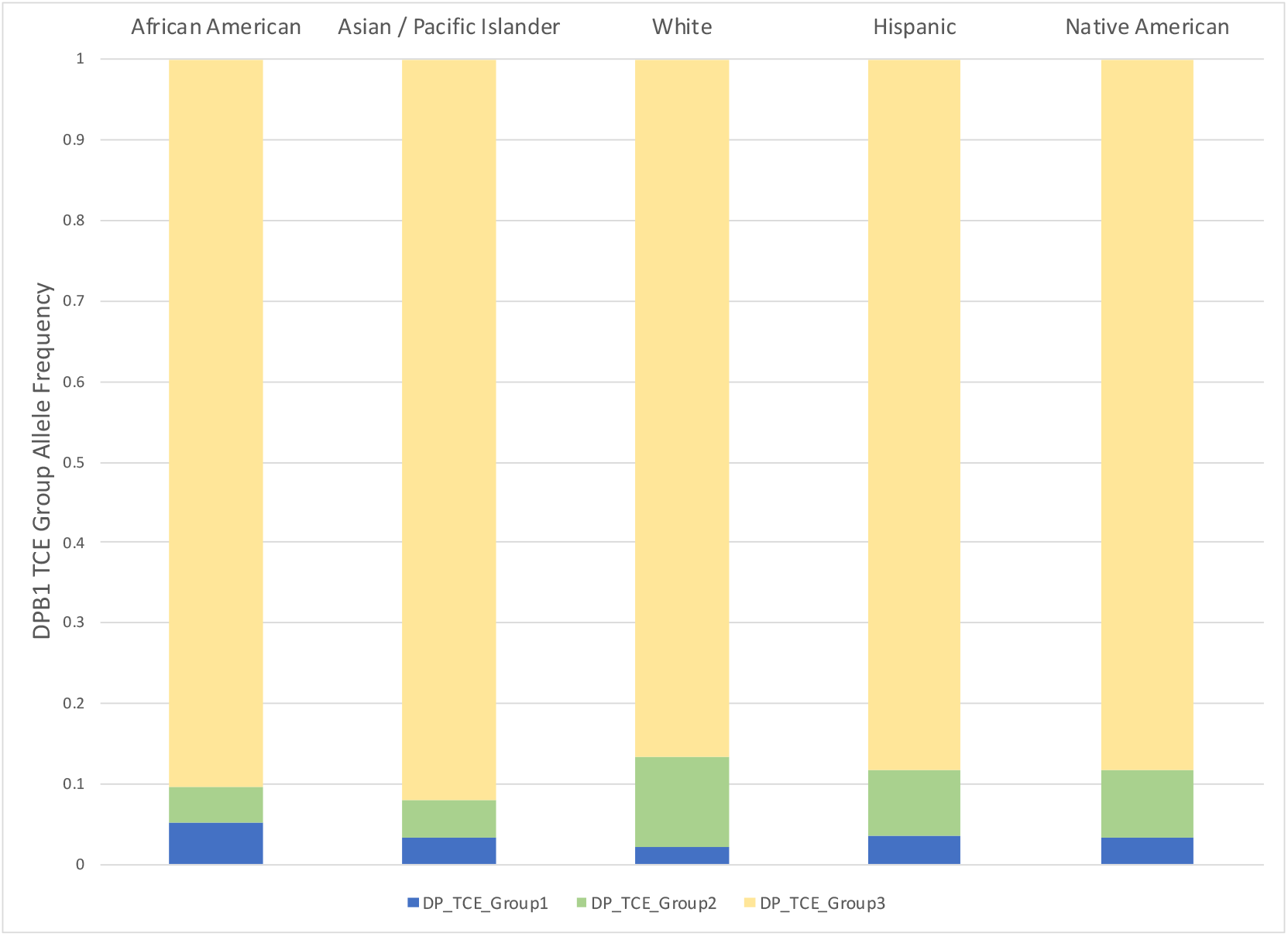
HLA-DPB1 frequencies at the T-Cell Epitope (TCE) group level within five US broad population categories. The DPB1 allele-to-TCE-group mappings are described by Crivello et al^29^. The alleles in these groups have been characterized as either immunogenic (Group 1), intermediately immunogenic (Group 2), or poorly immunogenic (Group 3). The frequency of DPB1 alleles that belong to each respective TCE category are summed together.

**Figure 4.**
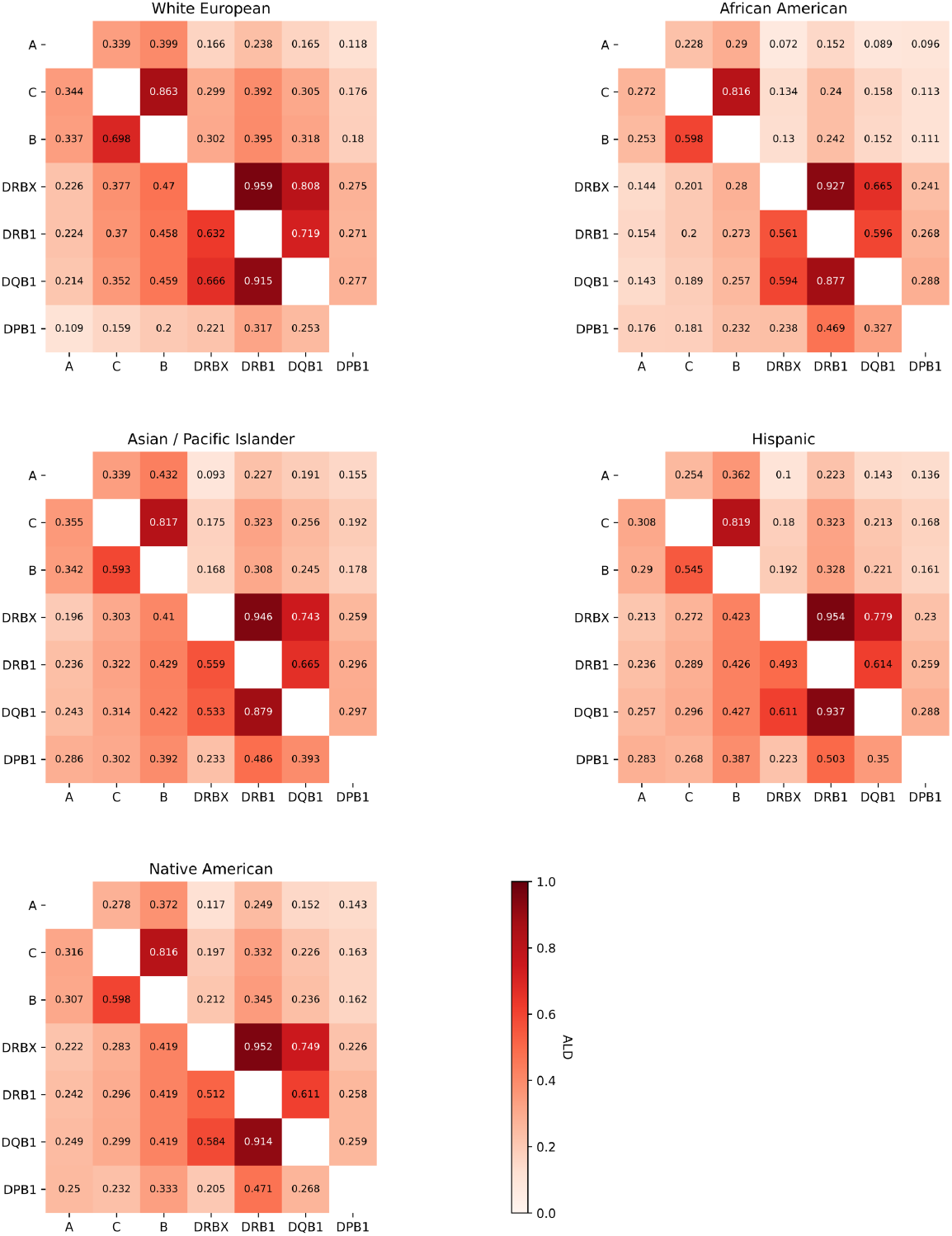
Asymmetric linkage disequilibrium (ALD) values between A, C, B, DRB3/4/5, DRB1, DQB1 and DPB1 loci within five US broad population categories. The values in each box represent how predictable the allele is for the row locus given knowledge of which HLA allele is present in the column locus. Asymmetry in mutual information is caused by HLA loci having differing numbers of alleles. The alleles for the DRB3, DRB4, DRB5 loci and the lack of any DRB3/4/5 gene are treated as a superlocus due to copy number variation, abbreviated with a four-character name “DRBX”.

**Figure 5.**
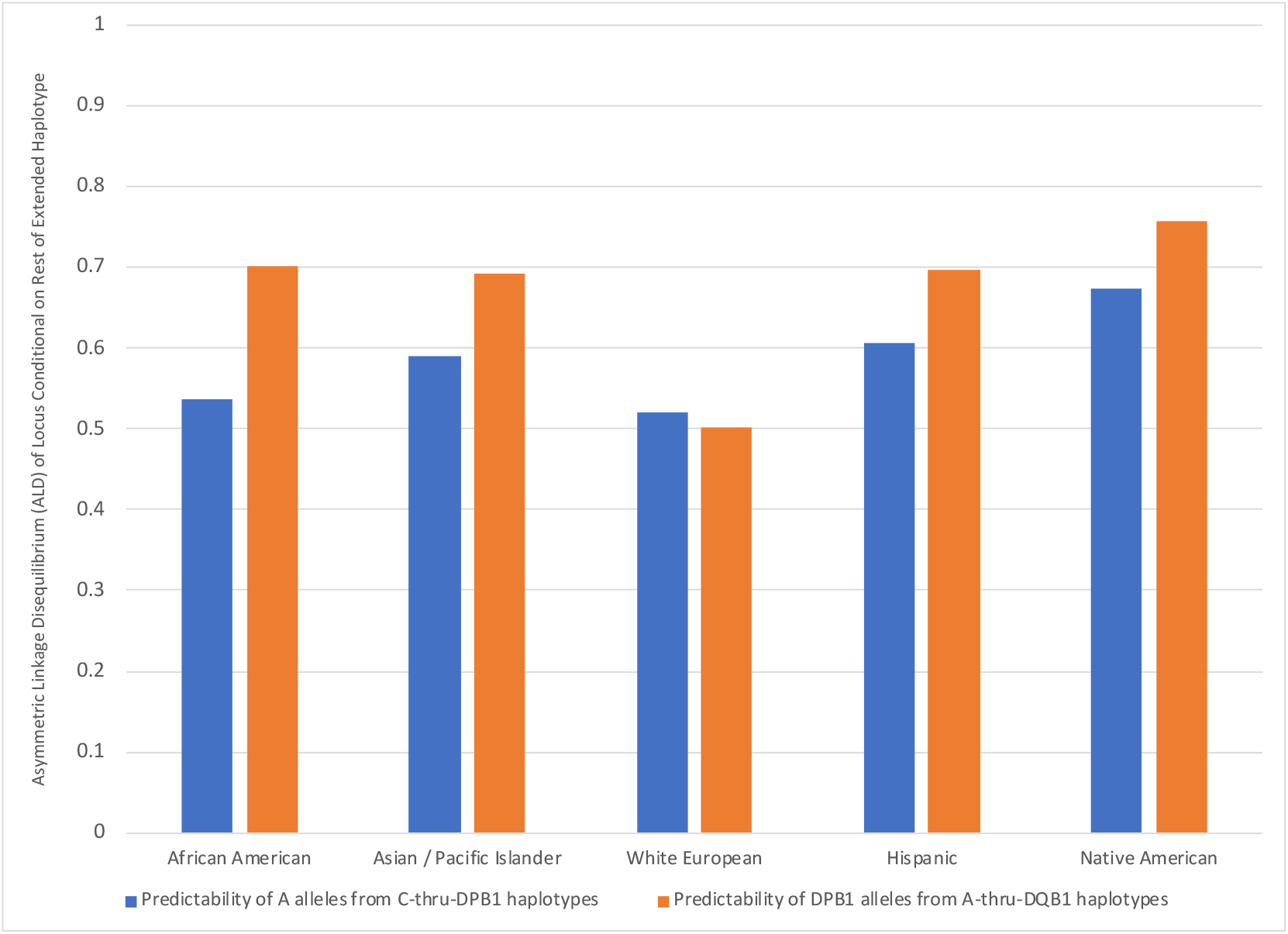
Predictability of missing HLA-DPB1 locus alleles versus missing HLA-A locus alleles using extended haplotype frequencies among 5 broad population categories. This asymmetric linkage disequilibrium (ALD) analysis shows the predictability of alleles at a single locus, conditional on the remainder of the extended haplotype block. We compare a scenario where C-thru-DPB1 haplotypes are used to predict a missing allele at the HLA-A locus (blue columns), versus when A-thru-DQB1 haplotypes used to predict a missing HLA-DPB1 allele (orange columns).

## DISCUSSION

In this study we found that most patients who have high resolution HLA-matched donors for the A, B, C, and DRB1 loci (8-of-8 matched) will also have an DP-TCE permissive donor in the US registry. Our match likelihood models considered both high resolution matching for DPB1 alleles and permissive mismatching for DP-TCE groups. In some circumstances, e.g. transplantation for non-malignant disease, DPB1 high resolution allele-matched donors may be preferred to a DP-TCE permissive mismatched donor because of the lower potential risk of graft-versus-host-disease with allele matching, with a tradeoff of higher risk of relapse. Based on modeling match likelihoods for both match stringencies, we determined, not surprisingly, that a DP-TCE permissibly mismatched donor is indeed relatively easier to find than a DPB1-matched donor because more than one common DPB1 allele falls into each T-cell epitope group.

Our study informs search strategies when a more optimally matched donor with DP-TCE permissibility is sought. In cases where a patient has several 8-of-8 matched donors without DPB1 typing, our HLA frequencies allowed us to compute for a given patient the expected number of donors where DPB1 confirmatory typing should be ordered before a more optimal DP-TCE permissibly mismatched donor is found.

Underlying the variation across patient populations, we found population differences in frequency of DPB1 alleles and DPB1 permissive categories. The self-identified race/ethnicity of the donor substantially alters the posterior probability of the distribution of possible DPB1 alleles, conditional on the alleles present at the other A, C, B, DRB1 loci. Our models for the number of donors need to type to find a DPB1 match are consistent with the lower bound found by random potentially-matched donor selection for confirmatory typing^17^. Our results further show that the HLA typing at other loci and self-identified race/ethnicity could be used operationally to prioritize donors for additional typing that are more likely to be matched at DPB1, reducing HLA typing costs and time-to-transplant.

We found that many extended HLA haplotypes with the same A-thru-DQB1 alleles, there was little allelic variation at DPB1, and that across population the known recombination hotspot between the DQ and DP gene complexes has had variable historical impacts on linkage disequilibrium. Among all populations studied, DPB1 alleles were the least predictable from the other loci in the European White population, the only population where DP recombination rates had been studied^28^, indicating that lack of consideration of race/ethnic diversity in immunogenetics studies has hindered our understanding of HLA matching. More research is needed to understand the evolutionary mechanisms driving conservation of HLA haplotype blocks and if these mechanisms differ across populations.

Our modeling results also overturn a commonly reported intuition that matching at DPB1 locus would be much less practical due to high allelic diversity at DPB1 and presence of a recombination hotspot that separates the DP gene complex from the rest of the HLA region. These inferences may have been flawed because they were based on misinterpretation of retrospective analyses of transplant pairs where a DPB1-matched donor was rarely sought.

The higher-than-expected predictability of DPB1 alleles provides rationale for development of the next generation of donor search algorithms that would make it much more practical for transplant centers to identify a more closely matched donor that would confer better outcomes for their patients. The capability to incorporate DPB1 prediction into the NMDP HapLogic matching algorithm has recently been implemented. Our results indicate that integrated DPB1 predictions will have great utility given the low rate of historical typing of DPB1 and models that show high operational search productivity when seeking DP-TCE permissibility.

The haplotype frequency estimates have some limitations in that they do not yet include all classical HLA loci. Matching at DRB3/4/5, DQA1 and DPA1 has not yet been shown independently to be clinically relevant for HSCT matching, although alleles at these loci are potential targets of patient antibody mediated rejection^29^. Mismatches across certain DQA1∼DQB1 heterodimers^26^ and low-expression loci^30^ may also increase risks. Typing across all classical HLA loci has occurred for new NMDP donor recruits only since 2020, thus the proportion of registry donors typed at the DQA1 and DPA1 loci remains very low (1.5%). Fortunately, because of high linkage disequilibrium between the DQA1 and DQB1 and the DPA1 and DPB1 genes respectively, the alpha chain alleles are likely to be highly predictable from the beta chain alleles that form the other half of the Class II heterodimer molecules based on very limited available two-locus haplotype data published to date^31,32^.

In summary, this study extends models for HCT unrelated donor registry matching to the DPB1 locus, providing valuable information for clinicians seeking a more optimal HLA-matched donor for their patients. The haplotype frequency data included in this report has expanded the loci predicted by the US registry match algorithm also has wide application for basic research in immunogenetics and association studies as well as clinical translational applications in transplantation and vaccine design.

## Data Availability

All data produced in the present study are available upon reasonable request to the authors

http://doi.org/10.5281/zenodo.4474442

## ACKNOWLEDGEMENTS

The bioinformatics methods used for this analysis were developed through a research grant funded by the US Office of Naval Research (N00014-20-1-2832). The CIBMTR is supported by grant 5U24-CA076518 from the National Institutes of Health, National Cancer Institute, National Heart, Lung, and Blood Institute, and National Institute of Allergy and Infectious Diseases; and by grant 5U10HL069294 from the National Institutes of Health, National Heart, Lung, and Blood Institute and National Cancer Institute. We thank the HLA typing labs for contributing the genotyping data to the NMDP registry. Special thanks to the 40+ million volunteer registry members around the world. We thank Kevin Tram for review of the study design. We thank Mike Halagan for contributing to a preliminary version of the study and Dr. Carolyn Hurley for input on the study design. This paper is dedicated to the memory of Dr. Robert J. Hartzman.

## AUTHORSHIP CONTRIBUTIONS

LG, SS, BS, and MM contributed to the study design and wrote the paper. LG and MM conducted the haplotype frequency analysis and ran the match likelihood models. All authors reviewed and revised the paper.

