## Supplemental Material for "Unrelated Stem Cell Donor HLA Match Likelihoods in the US Registry Incorporating HLA-DPB1 Permissive Mismatching"

**SUPPLEMENTAL DATA:**

**Supplementary Figure 1A. Historical adult donor match rates for 8-of-8 TCE match stringency with for 5 population categories.** This match stringency requires high resolution allele matching at the A, B, C, DRB1 loci and DPB1 TCE permissive or DPB1 high resolution matched. The 2015-2016 discontinuity was caused by the addition of international cooperative registries to NMDP upfront search, rather than organic recruitment of US donors.

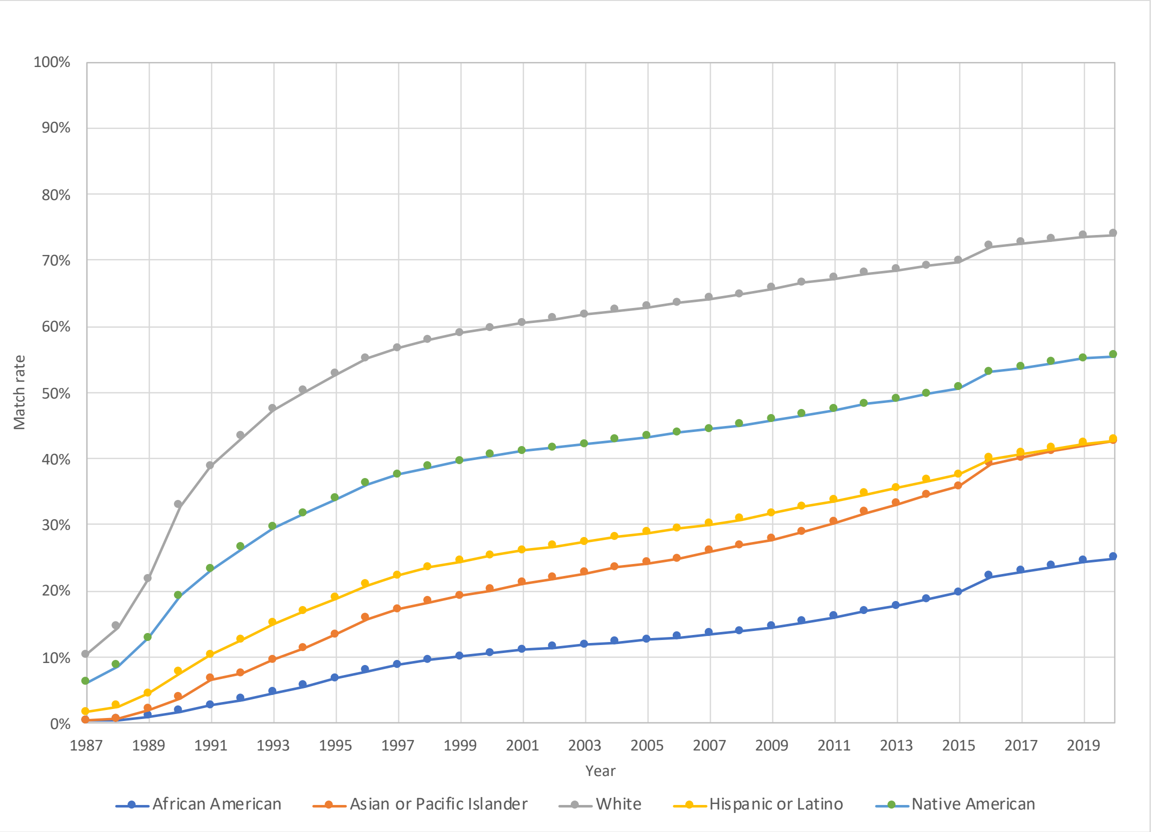

**Supplementary Figure 1B. Historical adult donor match rates for the 7-of-8 TCE matching stringency for 5 broad population categories.** This match stringency allows for up to one mismatch at any locus considering high resolution allele match at the A, B, C, DRB1 loci and DPB1 TCE permissive or DPB1 high resolution matched.

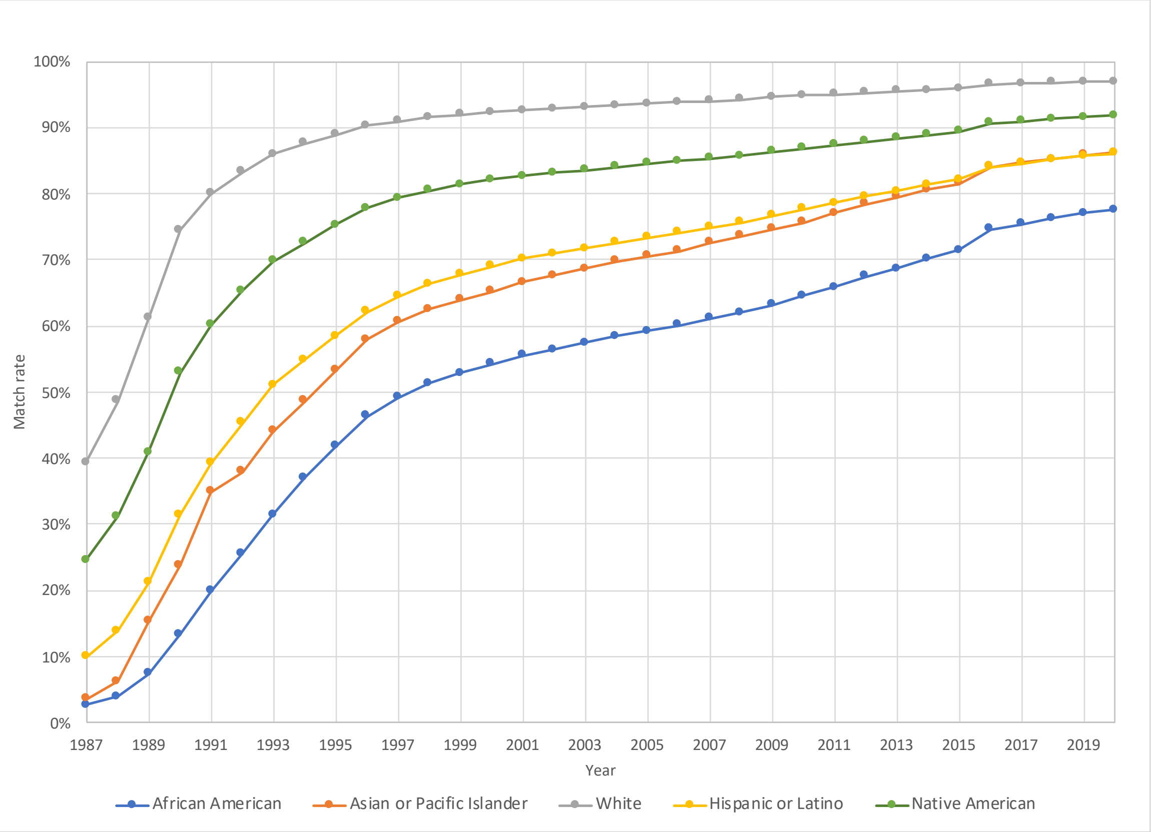

**Supplementary Figure 1C. Historical adult donor match rates for 10-of-10 TCE match stringency with for 5 population categories.** This match stringency requires high resolution allele matching at the A, B, C, DRB1, DQB1 loci and DPB1 TCE permissive or DPB1 high resolution matched.

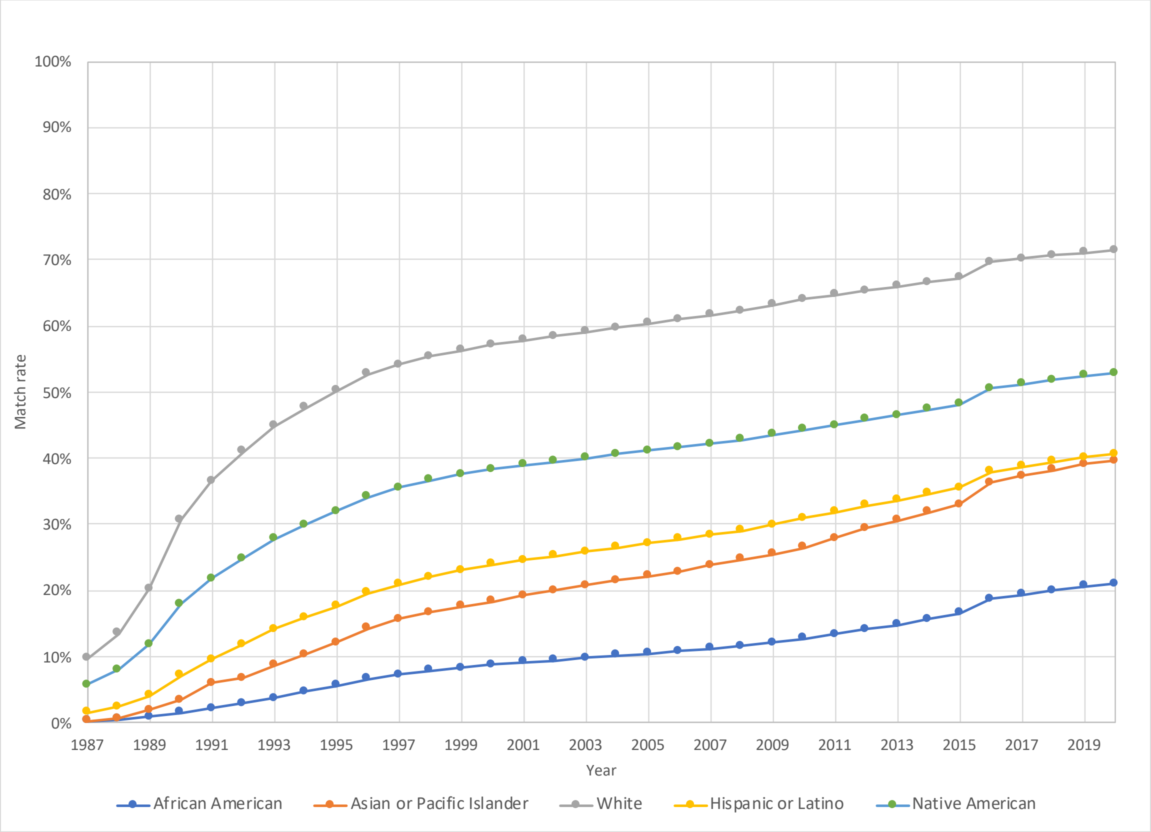

**Supplementary Figure 1D. Historical adult donor match rates for the 9-of-10 TCE matching stringency for 5 broad population categories.** This match stringency allows for up to one mismatch at any locus considering high resolution allele match at the A, B, C, DRB1, DQB1 loci and DPB1 TCE permissive or DPB1 high resolution matched.

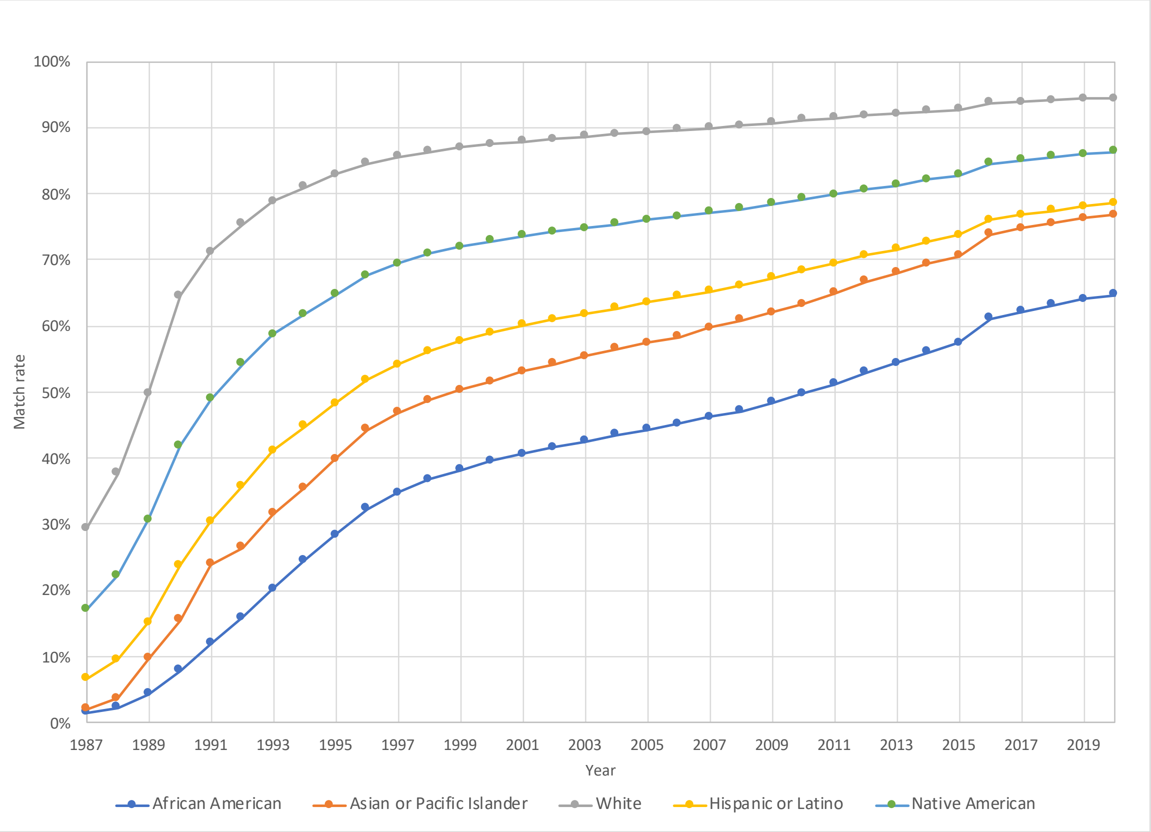

**Supplementary Figure 1E. Historical adult donor match rates for 12-of-12 match stringency with for 5 population categories.** This match stringency requires high resolution allele matching at the A, B, C, DRB1, DQB1, DPB1 loci.

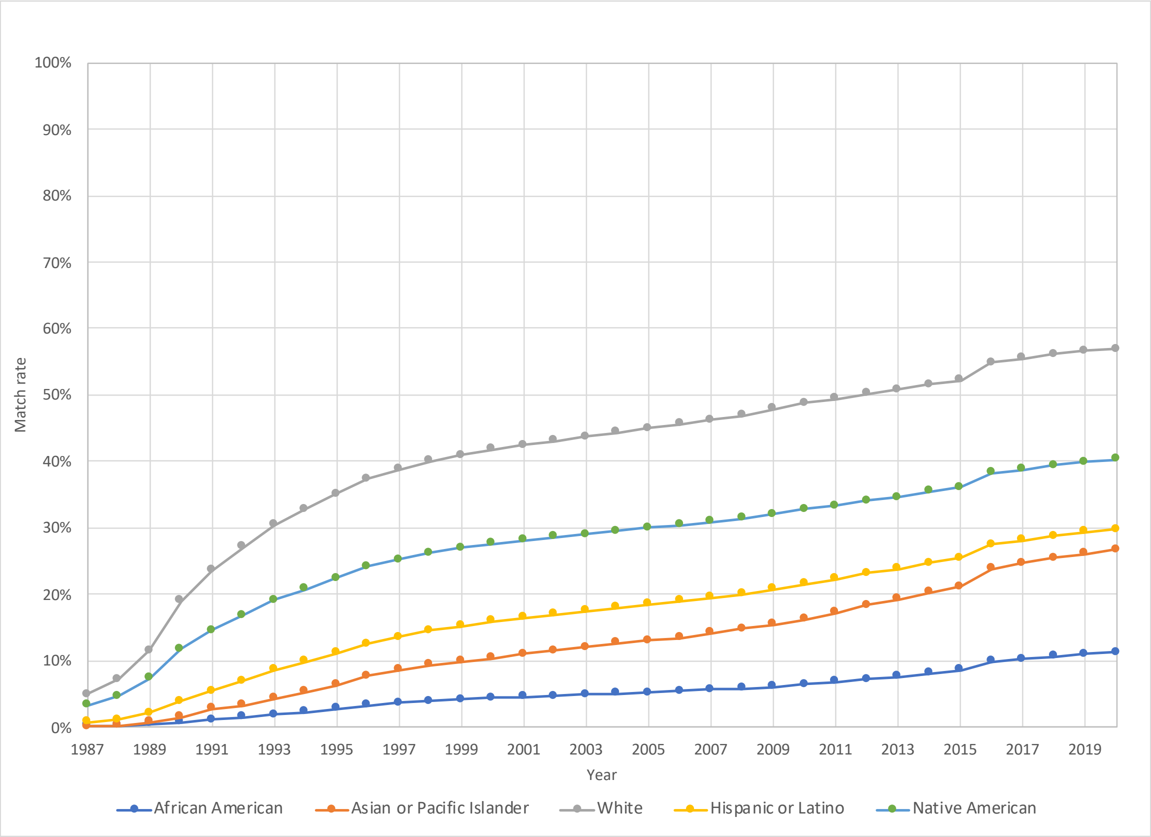

**Supplementary Figure 2A. Adult donor HLA match likelihoods for the US registry among 21 detailed population categories as of end of 2020.** The "8of8" match stringency requires high resolution allele matching at the HLA loci A, B, C, DRB1. The "8of8 TCE" match stringency requires the donor to be additionally DPB1 T-Cell Epitope (TCE) permissive or DPB1 high resolution matched. The "10of10" match stringency requires high resolution allele matching at the HLA loci A, B, C, DRB1, DQB1. The "10of10 TCE" match stringency requires the donor to be additionally DPB1 TCE permissive or DPB1 high resolution matched. The "12of12" match stringency requires high resolution allele matching at the HLA loci A, B, C, DRB1, DQB1, DPB1.

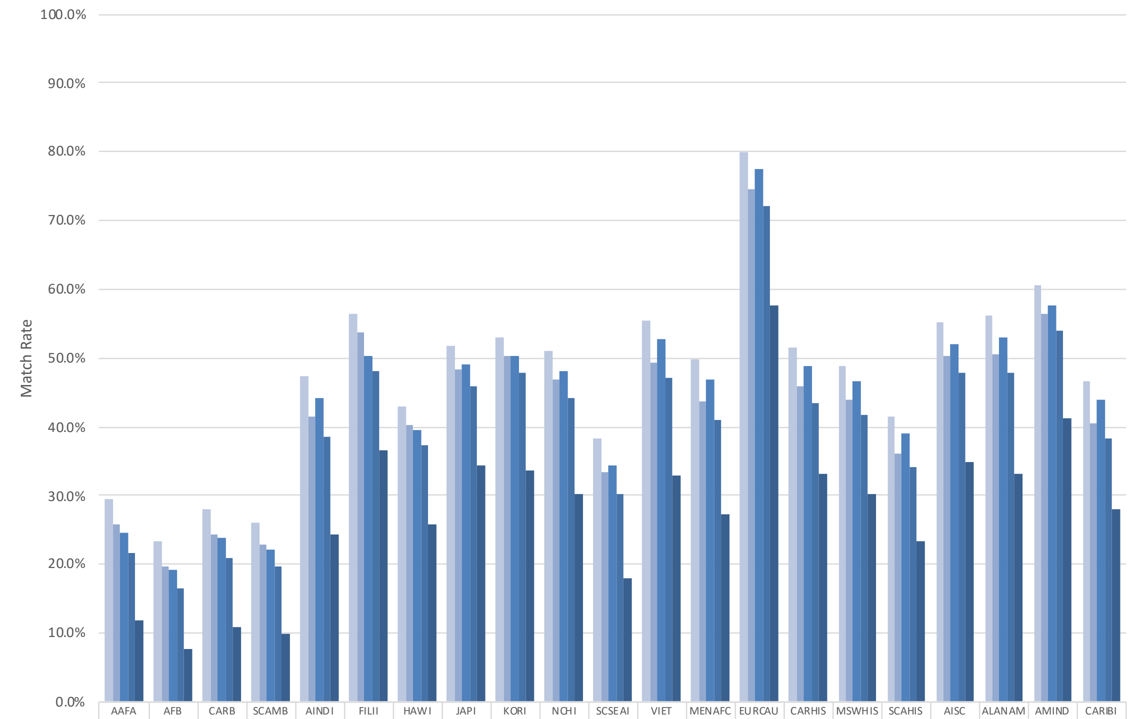


| | AAFA | AFB | CARB | SCAMB | AINDI | FILII | HAWI | JAPI | KORI | NCHI | SCSEAI | VIET | MENAFC | EURCAU | CARHIS | MSWHIS | SCAHIS | AISC | ALANAM | AMIND | CARIBI |
|---|---|---|---|---|---|---|---|---|---|---|---|---|---|---|---|---|---|---|---|---|---|
| 8of8 | 29.5% | 23.4% | 28.1% | 26.2% | 47.4% | 56.3% | 42.9% | 51.7% | 53.0% | 51.1% | 38.3% | 55.4% | 49.9% | 79.8% | 51.6% | 48.9% | 41.5% | 55.1% | 56.2% | 60.6% | 46.7% |
| 8of8 TCE | 25.8% | 19.7% | 24.4% | 22.8% | 41.6% | 53.8% | 40.4% | 48.2% | 50.2% | 46.8% | 33.4% | 49.3% | 43.8% | 74.5% | 45.9% | 43.8% | 36.2% | 50.4% | 50.6% | 56.5% | 40.5% |
| 10of10 | 24.5% | 19.2% | 23.9% | 22.2% | 44.1% | 50.3% | 39.5% | 49.1% | 50.3% | 48.2% | 34.4% | 52.9% | 46.8% | 77.4% | 48.8% | 46.7% | 39.0% | 52.1% | 53.0% | 57.7% | 43.8% |
| 10of10 TCE | 21.7% | 16.5% | 20.9% | 19.7% | 38.6% | 48.2% | 37.4% | 46.0% | 47.8% | 44.1% | 30.1% | 47.1% | 41.0% | 72.0% | 43.5% | 41.9% | 34.1% | 47.8% | 47.8% | 53.9% | 38.2% |
| 12of12 | 11.8% | 7.7% | 10.9% | 9.9% | 24.4% | 36.6% | 25.7% | 34.5% | 33.8% | 30.2% | 18.1% | 32.9% | 27.4% | 57.5% | 33.3% | 30.2% | 23.5% | 34.9% | 33.3% | 41.2% | 28.1% |

**Supplementary Figure 2B: Adult donor HLA match likelihoods for the US registry among 21 detailed population categories as of end of 2020, allowing for one mismatch at any locus.** The "7of8" match stringency allows for up to one high resolution allele mismatch at the HLA loci A, B, C, DRB1. The "7of8 TCE" match stringency additionally considers matching at DPB1 T-Cell Epitope (TCE) permissive or DPB1 high resolution allele, allowing for up to one mismatch. The "9of10" match stringency allows for up to one high resolution allele mismatch at the HLA loci A, B, C, DRB1, DQB1. The "9of10 TCE" match stringency additionally considers matching at DPB1 TCE permissive or DPB1 high resolution allele, allowing for up to one mismatch. The "12of12" match stringency allows for up to one high resolution allele mismatch at the HLA loci A, B, C, DRB1, DQB1, DPB1.

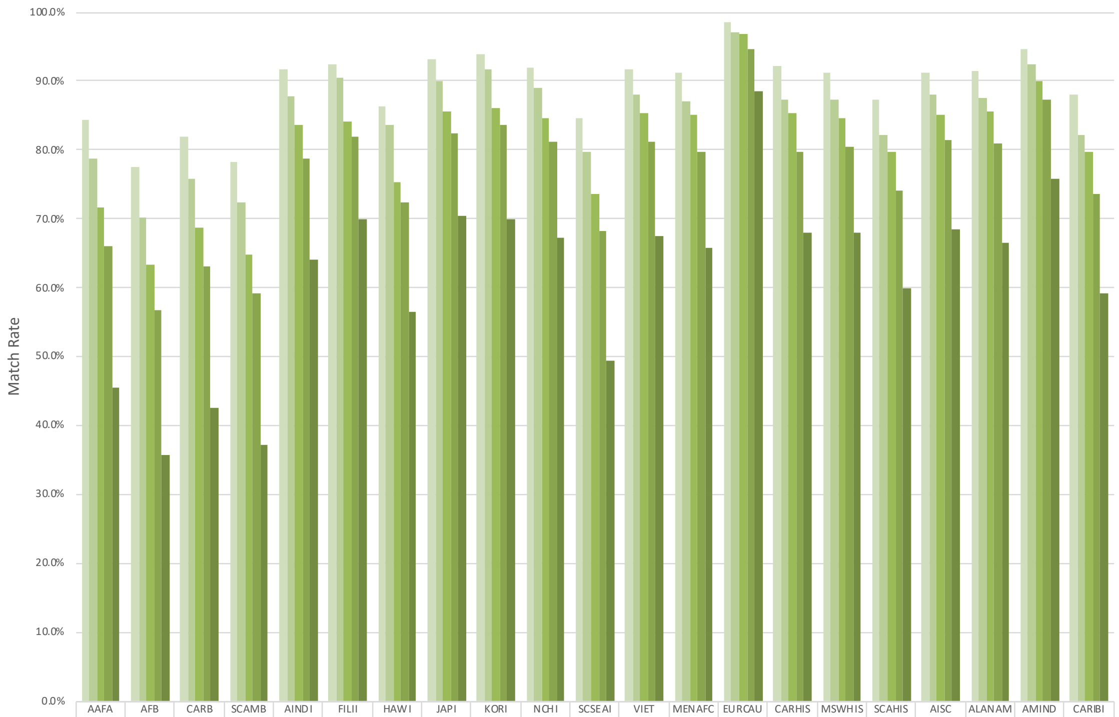


| | AAFA | AFB | CARB | SCAMB | AINDI | FILII | HAWI | JAPI | KORI | NCHI | SCSEAI |
|---|---|---|---|---|---|---|---|---|---|---|---|
| 7of8 | 84.4% | 77.5% | 82.0% | 78.2% | 91.7% | 92.5% | 86.2% | 93.1% | 93.8% | 91.8% | 84.7% |
| 7of8 TCE | 78.8% | 70.3% | 75.8% | 72.3% | 87.7% | 90.4% | 83.6% | 89.9% | 91.6% | 88.9% | 79.6% |
| 9of10 | 71.7% | 63.2% | 68.8% | 64.8% | 83.6% | 84.1% | 75.2% | 85.5% | 86.1% | 84.7% | 73.6% |
| 9of10 TCE | 66.1% | 56.8% | 63.0% | 59.2% | 78.8% | 82.0% | 72.4% | 82.3% | 83.7% | 81.2% | 68.2% |
| 11of12 | 45.6% | 35.7% | 42.6% | 37.3% | 64.0% | 69.8% | 56.5% | 70.4% | 69.9% | 67.2% | 49.5% |

| | VIET | MENAFC | EURCAU | CARHIS | MSWHIS | SCAHIS | AISC | ALANAM | AMIND | CARIBI |
|---|---|---|---|---|---|---|---|---|---|---|
| 7of8 | 91.6% | 91.2% | 98.6% | 92.2% | 91.1% | 87.3% | 91.1% | 91.4% | 94.6% | 88.1% |
| 7of8 TCE | 88.1% | 86.9% | 97.1% | 87.3% | 87.3% | 82.1% | 88.1% | 87.6% | 92.5% | 82.2% |
| 9of10 | 85.2% | 85.1% | 96.8% | 85.2% | 84.7% | 79.8% | 85.2% | 85.5% | 90.0% | 79.8% |
| 9of10 TCE | 81.2% | 79.8% | 94.7% | 79.8% | 80.3% | 74.1% | 81.4% | 80.8% | 87.2% | 73.6% |
| 11of12 | 67.5% | 65.7% | 88.6% | 68.0% | 68.0% | 60.0% | 68.6% | 66.6% | 75.7% | 59.2% |

**Supplementary Figure 3: Donors2Type among 21 detailed population categories.**

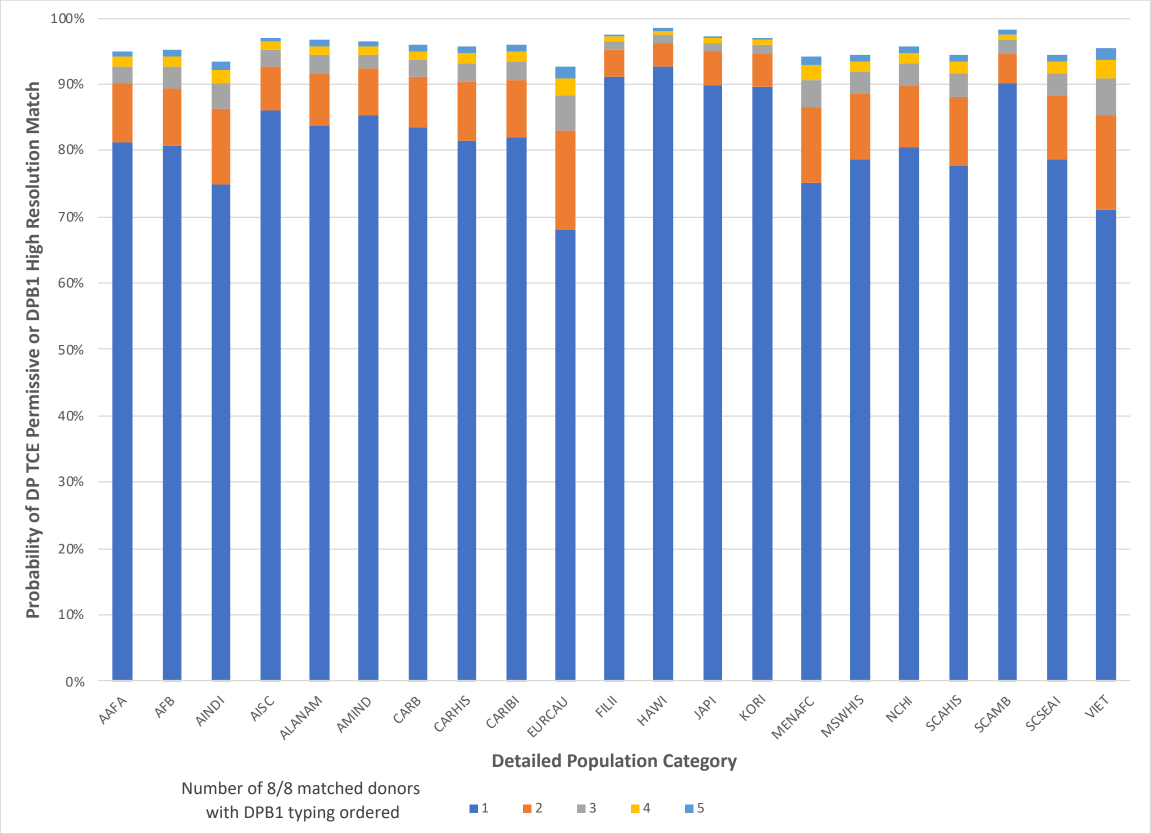

**Supplemental Table 1: Number of donors in the NMDP/Be The Match Registry at year-end from 1987 – 2020.** The NMDP registry started operation in 1987. This table provides the number of donors in that database at the end of each year of operation. The US donors are a subset of the total donors that reside in the United States. These number reflect increases due to recruitment and aggregation (international registries adding their donors to the NMDP database) and also decreases due to attrition from the removal of donors from the registry due to attrition (age 61), disinterest, inability to contact or medical deferral.

| Year | US Donors | Total Donors |
|---|---|---|
| 1987 | 11,261 | 11,261 |
| 1988 | 24,478 | 24,479 |
| 1989 | 69,710 | 72,947 |
| 1990 | 240,926 | 245,636 |
| 1991 | 470,801 | 477,316 |
| 1992 | 687,107 | 746,466 |
| 1993 | 932,003 | 1,125,623 |
| 1994 | 1,236,997 | 1,516,380 |
| 1995 | 1,529,325 | 1,974,973 |
| 1996 | 2,017,631 | 2,585,531 |
| 1997 | 2,370,139 | 3,034,934 |
| 1998 | 2,689,327 | 3,482,779 |
| 1999 | 2,954,098 | 3,848,181 |
| 2000 | 3,226,836 | 4,220,706 |
| 2001 | 3,464,497 | 4,557,886 |
| 2002 | 3,663,049 | 4,890,235 |
| 2003 | 3,825,197 | 5,232,123 |
| 2004 | 4,030,040 | 5,622,728 |
| 2005 | 4,220,585 | 5,980,250 |
| 2006 | 4,449,264 | 6,393,169 |
| 2007 | 4,824,476 | 6,874,852 |
| 2008 | 5,149,624 | 7,383,820 |
| 2009 | 5,617,764 | 8,139,295 |
| 2010 | 6,162,087 | 8,971,126 |
| 2011 | 6,602,736 | 9,755,035 |
| 2012 | 7,034,782 | 10,765,482 |
| 2013 | 7,379,343 | 11,613,510 |
| 2014 | 7,724,211 | 12,740,364 |
| 2015 | 8,038,996 | 13,853,755 |
| 2016 | 8,261,119 | 18,275,915 |
| 2017 | 8,490,239 | 19,560,679 |
| 2018 | 8,751,037 | 21,001,265 |
| 2019 | 8,846,348 | 22,377,119 |
| 2020 | 8,910,469 | 23,314,772 |

**SUPPLEMENTAL METHODS:**

**Modeling 8/8 with DP T-Cell Epitope (TCE) Group Permissive Mismatching:**

The first step in modeling DP TCE Matching is to map all HLA-DPB1 alleles to the 3 TCE groups. The DPB1 allele-to-TCE-group mappings are described by Crivello et al. The alleles in these groups have been characterized as either immunogenic (Group 1), intermediately immunogenic (Group 2), or poorly immunogenic (Group 3).

The allele frequencies for DP TCE groups in the 21 populations are given below in Supplementary Methods Table 1. The data is displayed

**Supplementary Methods Table 1: DP TCE Group Frequencies for 21 US Populations**

| POP | TCE1 | TCE2 | TCE3 |
|---|---|---|---|
| AAFA | 0.0833 | 0.0700 | 0.8468 |
| AFB | 0.0876 | 0.0928 | 0.8196 |
| AINDI | 0.0767 | 0.1289 | 0.7944 |
| AISC | 0.0602 | 0.1329 | 0.8069 |
| ALANAM | 0.0408 | 0.1323 | 0.8269 |
| AMIND | 0.0410 | 0.1351 | 0.8239 |
| CARB | 0.0807 | 0.0868 | 0.8325 |
| CARHIS | 0.0776 | 0.1521 | 0.7704 |
| CARIBI | 0.0802 | 0.1480 | 0.7719 |
| FILII | 0.0317 | 0.0582 | 0.9101 |
| HAWI | 0.0437 | 0.0998 | 0.8565 |
| JAPI | 0.0967 | 0.0919 | 0.8113 |
| KORI | 0.0496 | 0.0702 | 0.8802 |
| MENAFC | 0.0524 | 0.1302 | 0.8174 |
| MSWHIS | 0.0467 | 0.1403 | 0.8130 |
| EURCAU | 0.0379 | 0.1421 | 0.8199 |
| NCHI | 0.0363 | 0.1228 | 0.8409 |
| SCAHIS | 0.0600 | 0.1328 | 0.8072 |
| SCAMB | 0.0546 | 0.1302 | 0.8152 |
| SCSEAI | 0.0674 | 0.1216 | 0.8110 |
| VIET | 0.0192 | 0.1870 | 0.7938 |

The following chart adapted from Zino et al. illustrates the rules for permissive mismatching of HLA-DP TCE genotypes.

**Supplementary Methods Figure 1: DP TCE Genotype Permissive Mismatch Algorithm:**

<table>
<tr><td rowspan="2" colspan="2"></td><th colspan="6">Recipient</th></tr>
<tr><th>1+1</th><th>1+2</th><th>1+3</th><th>2+2</th><th>2+3</th><th>3+3</th></tr>
<tr><td rowspan="6">Donor</td><td>1+1</td><td rowspan="3" colspan="3">Permissive</td><td rowspan="3" colspan="3">Non-Permissive (HvG)</td></tr>
<tr><td>1+2</td></tr>
<tr><td>1+3</td></tr>
<tr><td>2+2</td><td rowspan="3" colspan="3">Non-Permissive (GvH)</td><td rowspan="2" colspan="2">Permissive</td><td rowspan="2"></td></tr>
<tr><td>2+3</td></tr>
<tr><td>3+3</td><td colspan="2"></td><td>Permissive</td></tr>
</table>

This chart indicates that there are 6 possible genotypes for donors along the y-axis, and 6 possible DP TCE genotypes for recipients along the x-axis, for a total of 36 genotype pairs.

By running a 6-locus registry match likelihood model with 3 HLA-DP TCE group specificities along with the 10/10 A, C, B, DRB1, DQB1 allele specificities, the match probabilities where both TCE groups are matched along the diagonal was calculated.

However, there are additional 8 DP TCE donor-recipient genotype pairs that are permissive, but don't match at the DP TCE group level (1+1 with 1+2, 1+1 with 1+3, 1+2 with 1+1, 1+2 with 1+3, 1+3 with 1+1, 1+3 with 1+2, 2+2 with 2+3, 2+3 with 2+2) that aren't accounted for in the popchart model, which only gives the result for 6 matched genotypes along the diagonal of Supplementary Figure 1 (1+1 with 1+1, 1+2 with 1+2,1+3 with 1+3, 2+2 with 2+2, 2+3 with 2+3, 3+3 with 3+3).

Therefore, we needed additional calculations to measure the impact of those 8 boxes. The difference between the 10/10 and 10/10-with-DP-TCE is accounted for by the 30 genotype pairs that aren't matched for the TCE groups. 22 of those genotype pairs are non-permissive (shaded), and 8 are permissive but don't match. We next calculated the DP TCE group genotype pair frequencies for all 36 combinations in Supplementary Figure 1. The difference in match likelihood between 10/10-with-DP-TCE-Permissive and 10/10 is proportional to the DP TCE genotype pair frequency of the 22 non-permissive genotype pair boxes. The distance between 10/10 with DP TCE Permissive and 10/10 with DP TCE Exact Group Matching is proportional to the DP TCE genotype pair frequency of the 8 permissive genotype pair boxes.

The formula for solving for the 10/10-with-DP-TCE-Permissive-Mismatch match likelihood is given below:

Prob(10of10_with_DP_TCE_Permissive) = Prob(10of10) - ((Prob(10of10) - Prob(10of10_with_TCE_Exact_Group_Match)) * ((sum of 22 non-permissive genotype pair frequencies) / (sum of 30 mismatched genotype pair frequencies)))

Because TCE Group 3 is of high frequency, the final DP TCE Permissive match likelihoods are much closer to Prob(10of10_with_TCE_Exact_Group_Match) than to Prob(10of10).
